# Forecasting Ultra-early Intensive Care Strain from COVID-19 in England, v1.1.4

**DOI:** 10.1101/2020.03.19.20039057

**Authors:** Jacob Deasy, Emma Rocheteau, Katharina Kohler, Daniel J. Stubbs, Pietro Barbiero, Pietro Liò, Ari Ercole

## Abstract

The COVID-19 pandemic has led to unprecedented strain on intensive care unit (ICU) admission in parts of the world. Strategies to create surge ICU capacity require complex local and national service reconfiguration and reduction or cancellation of elective activity. These measures have an inevitable lag-time before additional capacity comes on-line. An accurate short-range forecast would be helpful in guiding such difficult, costly, and ethically challenging decisions.

At the time this work began, cases in England were starting to increase. If this represents a true spread in disease then ICU demand could increase rapidly. Here we present a short-range forecast based on published real-time COVID-19 case data from the seven National Health Service (NHS) commissioning regions in England (East of England, London, Midlands, North East and Yorkshire, North West, South East and South West). We use a Monte Carlo approach to model the likely impact of current diagnoses on regional ICU capacity over a 14-day horizon under the assumption that the increase in cases represents the start of an exponential growth in infections. Our model is designed to be parsimonious and based on available epidemiological data from the literature at the moment.

On the basis of the modelling assumptions made, ICU occupancy is likely to increase dramatically in the days following the time of modelling. If the current exponential growth continues, case numbers will be comparable to current ICU bed numbers within weeks. Despite variable growth in absolute patients, all commissioning regions are forecast to be heavily burdened under the assumptions used.

Whilst, like any forecast model, there remain uncertainties both in terms of model specification and robust epidemiological data in this early prospective phase, it would seem that surge capacity will be required in the very near future. Our findings should be interpreted with caution, but we hope that our model will help policy decision makers with their preparations. The uncertainties in the data highlight the urgent need for ongoing real-time surveillance to allow forecasts to be constantly updated using high quality local patient-facing data as it emerges.

## Introduction

The COVID-19 pandemic has rapidly caused an enormous worldwide medical and socioeconomic impact since the first case emerged on November 16th 2019 [1]. Although a self-limiting illness for most, the percentage of COVID-19 patients with hypoxaemic respiratory failure requiring ICU admission for mechanical ventilation translates into large numbers which may challenge healtcare provision. In Northern Italy, an exponential increase in COVID-19 admissions rapidly overwhelmed normal ICU capacity [2] and surge capacity had to be created quickly. The exact reason for the sudden need for ICU surge capacity in Italy, and whether this will generalise to other countries, is unclear, but both demographic factors and healthcare system structure are likely to be important. Notably, UK availability of ICU beds per capita compares poorly with other high-income countries—including Italy [3]. Whilst standard acute wards may be re-purposed easily, creating ICU capacity is constrained by the need for complex equipment and the delivery of highly specialised medical and nursing care. Nevertheless, there are mechanisms by which ICU capacity could be increased in an emergency, including facilitating ICU discharge of recovering patients through liberating downstream beds by reducing elective work, by stopping elective work likely to require ICU admission, or by changing referral policies. Such changes are likely to be relatively quick to implement but have important repercussions for normal healthcare provision and ideally would not be instituted until absolutely necessary. Furthermore, since ICUs in the UK typically undertake a high proportion of emergency work, much of which will continue despite the pandemic, and since occupancy is typically well above 80%, such strategies are likely to result in only a relatively modest increase in capacity. A greater increase in emergency physical capacity for mechanical ventilation could be achieved by opening new level 3 beds with additional equipment (e.g. operating theatre ventilators). This requires significant changes to infrastructure, processes, or staffing and is therefore logistically complex, expensive, and most importantly slow to implement. Forecasting was therefore essential in guiding such difficult policy decisions in Italy [2]. The explosion of cases seen in Italy means that an early warning of need for surge capacity is likely to be required in other countries including England. Epidemiological simulation has previously been successful in predicting the need for surge H1N1 ICU capacity in 2009 [4, 5]. In recent days, a similar simulation model for COVID-19 has been described [6], which suggests an overwhelming demand for critical care, with a peak occurring between May and early June 2020 and lasting 2 to 3 months depending on non-pharmacological intervention (NPI) assumptions. Such models are very useful but do not incorporate up-to-date data. Such projected timescales are unlikely to be reliable and are unsuitable for real-time surveillance and early warning. In this paper we use published COVID-19 diagnosis data for England to generate the earliest possible estimates of additional ICU demand due to infections in the coming days, based on cautious epidemiological data from the literature and *under the assumption that the current increase in cases represents the exponential phase of an outbreak* rather than a change in ascertainment. Our emphasis is on making an updatable model from the little time-series data that are available in this ultra-early period, with the understanding that assumptions are necessary where data are unavailable. Our model predictions use the latest results from the rapidly developing COVID-19 literature, account for English demographics, and are stratified by the National Health Service (NHS) commissioning regions across the country.

## Methods

We used COVID-19 diagnoses from England as reported by Public Health England (PHE) and matched to NHS commissioning regions [8] as our source data to obtain information on daily cases (reproduced in Figure 1). We started to extract this data feed on 13/03/20 to give daily case data.

**Figure 1:**
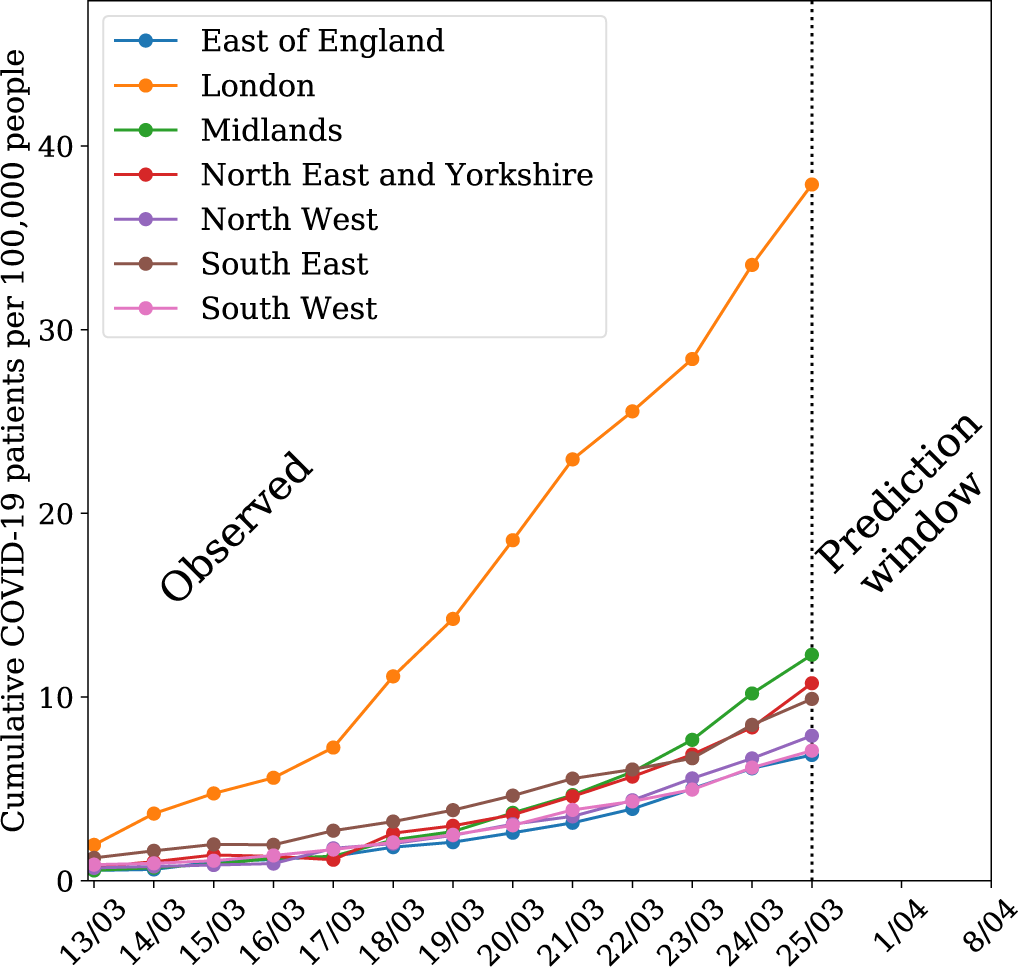
Model timeline. Our model relies on a *n*-day window for regression, beginning 13/03 with recent observed data. Themodel predicts two weeks into the future from the time of writing.

We assumed that the daily incidence of COVID-19 can be modelled as an exponential growth (in line with what was observed in Italy [2]). Therefore, we forecast the likely distributions of new COVID-19 diagnoses over the next 14 days by using an ordinary least squares fit to linearly extrapolate from the logarithm of the cumulative cases. We differentiated the cumulative model to obtain a daily incidence model i.e. we multiply by the exponent 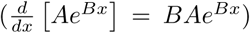, which stabilises the exponent fitting (*B*).

Using early data from Verity *et al*. 2020 [9] (reproduced in Table 1), we estimated the ICU admission rate by standardising to the local population in each NHS commissioning region in England (see Figure 2, obtained from the Clinical Commissioning Group population estimates for mid-2018 [10]). We adjust by age and sex, which have both been shown to strongly correlate with mortality [9]. The higher male:female ratio in China, according to data from the 2017 Chinese census [11], required us to adjust our calculations under the assumption that the added risk of being male holds on a univariate basis. We used the ICU admission rate estimates to obtain ICU admissions from the case predictions in each region.

**Table 1:** Mortality and critical care needs due to COVID-19 in England stratified by region and shown alongside ICU capacity (Data Sources: [6, 7]).

| Region | Mortality (%) | Require ICU (%) | No. Beds |
| --- | --- | --- | --- |
| East of England | 1.27 | 2.62 | 337 |
| London | 0.82 | 1.70 | 1019 |
| Midlands | 1.22 | 2.52 | 680 |
| North East and Yorkshire | 1.24 | 2.56 | 622 |
| North West | 1.19 | 2.46 | 643 |
| South East | 1.27 | 2.63 | 523 |
| South West | 1.41 | 2.92 | 303 |

**Figure 2:**
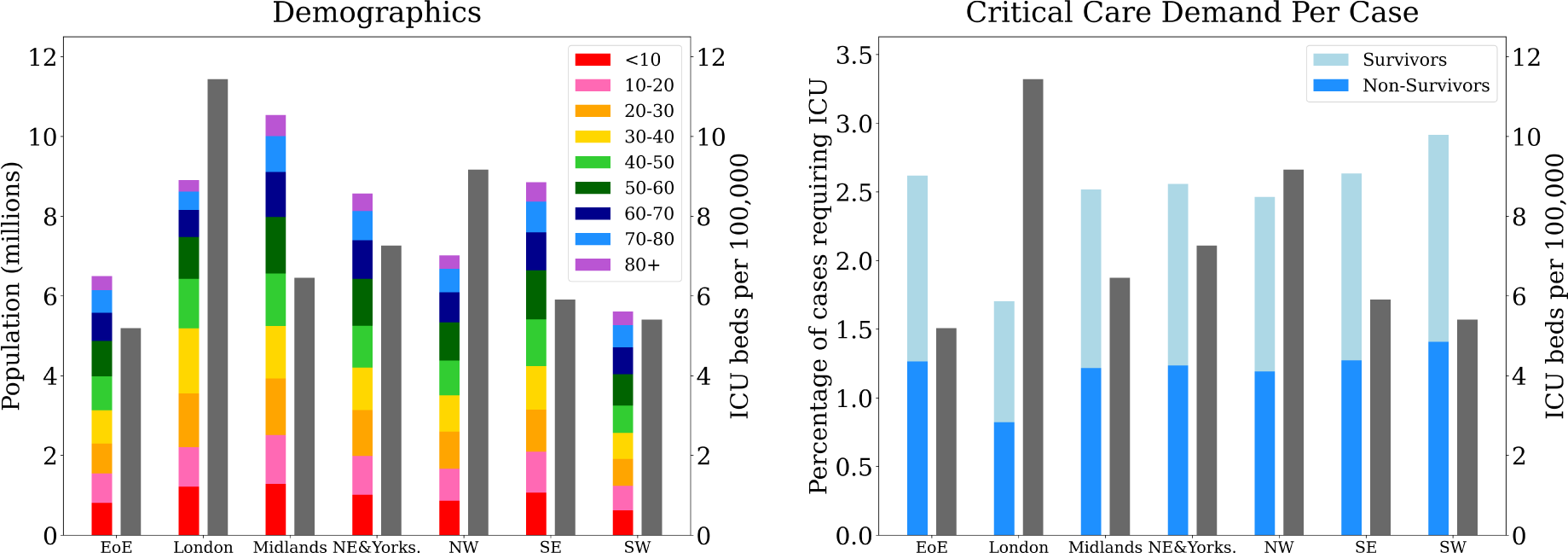
Regional demographics and expected critical care demand per case of COVID-19, stratified by region and compared to ICU bed capacity per 100,000 people. Population is divided into age categories and percentage of cases requiring ICU is divided into expected percentage of survivors and non-survivors. The numerical data can be seen in Table 1.

As new diagnoses have a delay to arrival in the ICU, we modelled this with a normal distribution (*µ* = 2, *(σ*= 3.5), allowing negative values as an indication of diagnosis after admission. The parameters of our delay distribution were based on multiple early reports from China [12, 13, 14, 15], which indicated a consistent pattern of disease progression: hospital admission occurring around day 8, acute respiratory distress syndrome (ARDS) on day 9, and admission to the ICU for mechanical ventilation on day 10. The strict requirements for testing in the UK would warrant hospitalisation in most cases [16], so we assumed cases to be diagnosed around the time of hospital admission, making the time to ICU around 2 days.

Predicting bed occupancy also required estimation of the length of stay for patients admitted to the ICU. The most up-to-date information indicates a median length of stay of approximately 8 days, with a wide interquartile range and positive skew [13, 14]. We modelled this with a gamma distribution (*α* = 8, β = 1).

To link the expected daily incidence to the expected ICU bed occupancy due to COVID-19 we used a Monte Carlo simulation with 500 samples. This involved sampling from the daily incidence model, as well as the delay and length of stay distributions to obtain bed occupancy along with respective uncertainty estimates. We represented bed occupancy as a percentage of the total number of ICU beds in the commissioning region based on data from a snapshot in December 2019 [7].

## Results

Figure 3 shows the expected number of cumulative COVID-19 patients assuming the current exponential trend in England. We found that the daily case growth in England was well fit by a linear model in log-space and therefore an exponential model for cumulative incidence. In the Midlands and the South West the fit is particularly good and the 95% confidence interval is tight, demonstrating these regions are likely to already be in a period of exponential growth of new cases. The fit is less reliable in the North East and Yorkshire, a region where we have separately confirmed the trend of observed new cases to have the weakest correlation with the other regions. The log-linear fit produces a strong *R*^2^ value in populous areas, strengthening the argument for exponential growth in areas where the virus has gained a foothold. Irrespective of how many of these cases translate into ICU patients, the projections indicate at least 100 cases per 100,000 people in all NHS commissioning regions in England within two weeks of writing.

**Figure 3:**
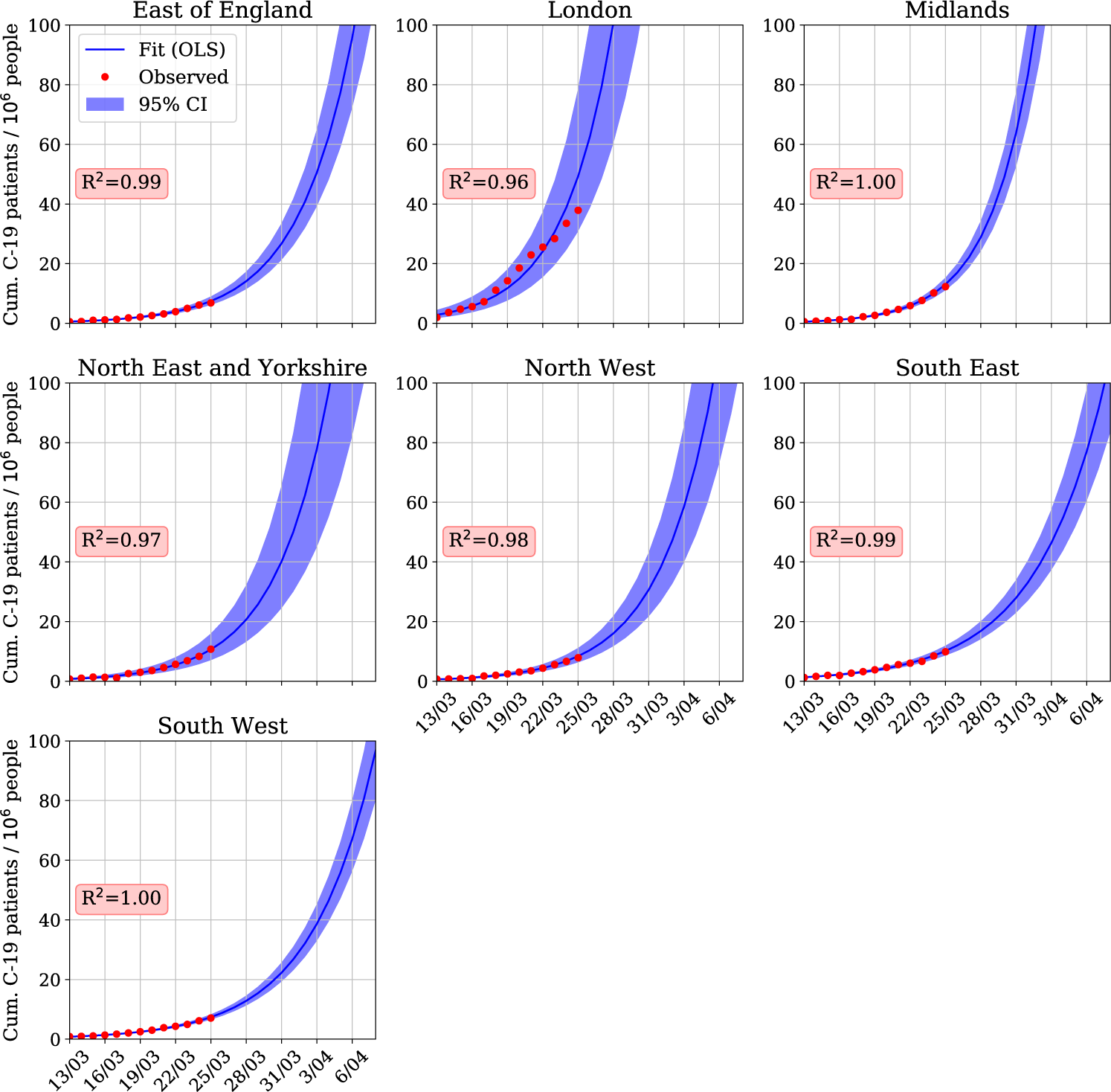
Projected cumulative COVID-19 patients per 100,000 people in the seven National Health Service commissioning regions in England. Exponential projections were calculated by an ordinary least squares fit in log-space to recent reported values. The figure shows the rapid expected increase in the number of COVID-19 patients, particularly in London and the Midlands.

Figure 4 shows the projected ICU occupancy due to COVID-19 from our model over a 14-day horizon for each of the NHS commissioning regions as a percentage of total ICU beds. The percentages reflect bed occupancy in addition to the normal cases. Notably, the confidence intervals here are tighter than the case projections in Figure 3 because ICU occupancy is a function of the past cases, and we have greater confidence in predicting the immediate future.

**Figure 4:**
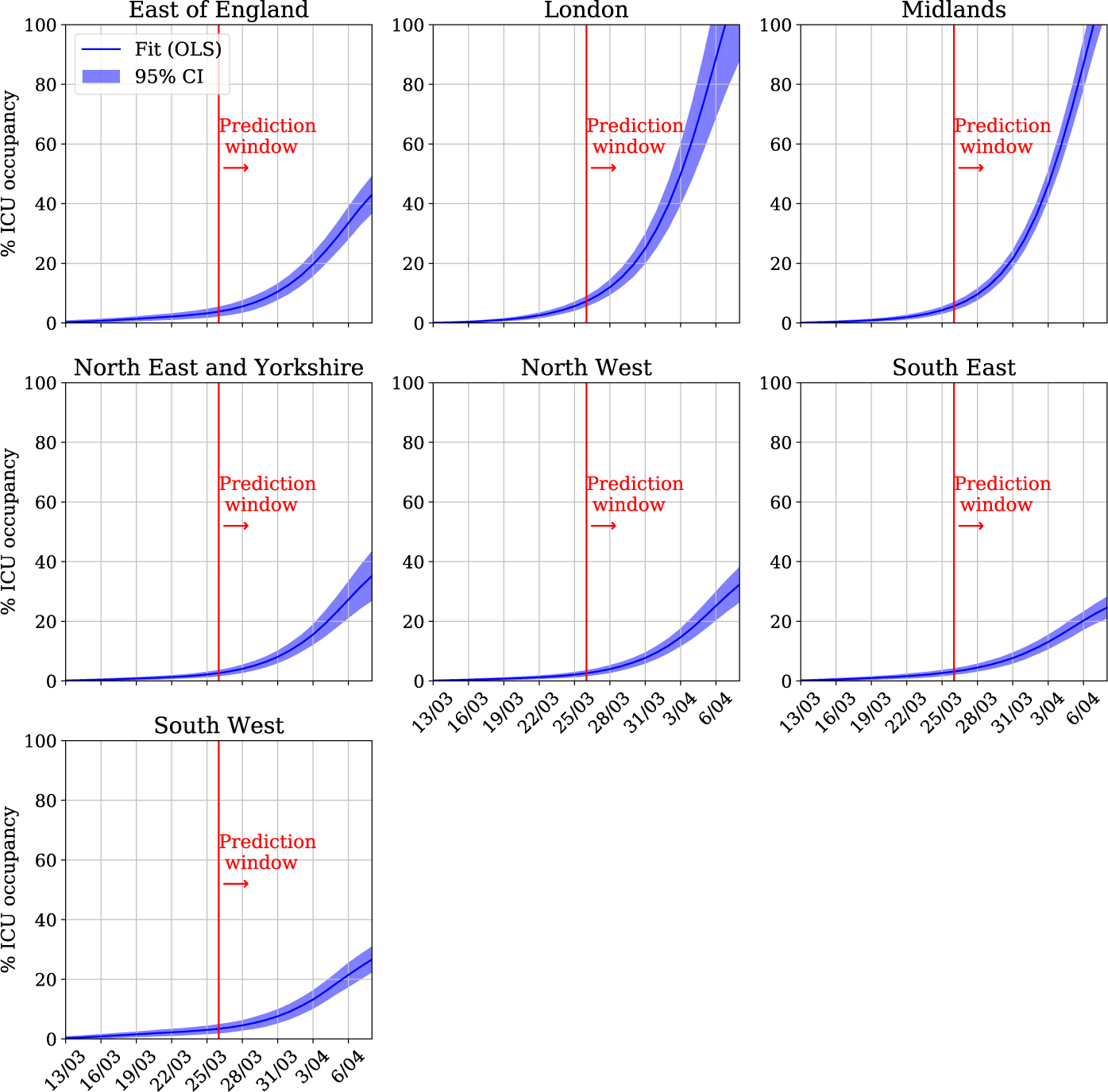
Projected regional COVID-19 ICU occupancy as a percentage of regional capacity in the seven National Health Service commissioning regions in England. The figure illustrates the large regional variation in projected ICU occupancy, with London and the Midlands reaching near 100% additional capacity due to COVID-19 within 14 days, whereas areas such as the South West and South East will only have approximately 20% of their beds occupied by COVID-19 patients.

Overall, Figure 4 shows predictions that will challenge ICU capacity in all areas, particularly in populous regions. London and the Midlands will gain a number of COVID-19 patients equivalent to 100% standard capacity within 14 days for slightly different reasons. In London, despite the higher number of ICU beds per capita, the higher case incidence per capita will overwhelm ICU capacity. Whereas in the Midlands, due to an increased average age, a higher percentage of ICU admissions is predicted (see Table 1), which combined with a higher number of cases than other regions will lead to exhausted capacity. As normal occupancy is often above 80%, even the additional minimum capacity due to COVID-19 of 20% in other regions will prove to be difficult to accommodate without capacity planning, to both increase the ICU bed availability and reduce the number of patients without COVID-19 requiring ICU admission.

All source code has been made available at https://github.com/ariercole/Cambridge_COVID-19_ICU [17]. In addition to our analysis and open-source code, an interactive model with current data is available at http://covid19icu.cl.cam.ac.uk. As documented cases in the literature evolve, we hope clinical and policy decision makers will be able to experiment based on their region or the statistics demonstrated by their cohort. Given the highly dynamic situation at the time this work was carried out, with model data changing on a daily basis, minimising model development time was crucial. Using agile project management methodologies, we were able to develop a working model, documentation, and web-implementation in less than one week.

## Discussion

In this study we demonstrate how publicly available data can be rapidly combined to dynamically model short-term ICU requirements in light of the emerging COVID-19 pandemic. Our data suggests that traditional ICU capacity could be rapidly consumed over a period of approximately 14 days from the time of modelling/writing, such figures hide substantial regional heterogeneity, with London and the Midlands demonstrating the most rapid growth. The key message from our paper varies with the temporal position of the reader. Within the current acute stage of this pandemic, we hope our findings provide a framework to facilitate the rapid development and deployment of additional ICU surge capacity as was required within days of the Italian pandemic [2]. In the future, we feel that our methodological approach (use of emerging epidemic curve data, Monte Carlo simulation, and population standardisation), as well as our simultaneous deployment of an interactive and dynamically updated web-tool have merit in modelling and communicating any future dynamically emerging event.

However, we do recognise that as with all models our is subject to significant assumptions and limitations. Both model and parameter uncertainties are inevitable, particularly when predicting the behaviour of a novel virus in a new population, and this may radically affect our forecasts. Nevertheless, we set out to provide the earliest possible data-driven forecast and therefore explicitly accept the limitations of the data that were available to us at the time. Our approach has been to keep the model as parsimonious as possible, with parameter estimates from the existing literature, to give a rough guide to early surge needs. With the small amount of available data, there are some specific limitations which need to be discussed. Our use of an updated, online resource we hope is a novel way of allowing interested parties to interact with the mathematical assumptions and formulate their own opinions.

We have used PHE published data for case ascertainment. We recognise that this data is potentially flawed and does not recognise all cases within the wider population. However, being as our explicit aim is to model the most severe cases, the effect of any ascertainment bias on our findings should be minimised due to the rollout of routine testing for all critical care patients as part of the “ COVID Hospitalisation in England Surveillance System—CHESS” [18]. Despite this, case ascertainment and delay between symptom onset and ICU admission is unlikely to be uniform between populations, healthcare settings, or discrete time points. Specifically, at the time of writing, test capacity for COVID-19 in the UK has led to stringent requirements for testing with only cases requiring hospitalisation being routinely tested [16]. By incorporating a ‘delay to ICU’ parameter within our model, that can take on negative values (thus indicating a diagnosis **after** ICU admission), we hope to have replicated the anecdotal experience of clinicians in these early phases of the pandemic; that a substantial proportion of patients are being confirmed as COVID-19 positive after ICU admission for respiratory distress. Our choice of this modelling assumption is based on not only on the clinical course of the disease but also aims to replicate the distortion of this natural history by current UK testing strategy. Again, to reflect the dynamism of this situation, we have allowed the user to alter this assumption via our online tool as the situation evolves.

Any flaw in our modelling assumptions can be considered as either rendering our model overly optimistic or pessimistic. Both have serious implications; optimism could cause detrimental delay in the development of surge capacity, whilst pessimism could cause substantial ‘knock-on’ morbidity (e.g. by prematurely cancelling elective work) as well as financial and system strain. Changes in WHO case-definition and surveillance strategies since the initial reporting of the pandemic mean that the original epidemiological data may overestimate ICU admission rates as milder community cases were not initially included. This would artificially inflate our forecast numbers as these data inform our standardised estimates of ICU cases from within each population [9]. Similarly, as already intimated, UK case definition has evolved over time. If the upturn in COVID-19 cases seen at the time of writing is largely driven by increased or altered ascertainment rather than a true rise in cases, then this would render our model overly pessimistic. Nevertheless, this will always be a limitation of any early modelling [9].

Although we have resolved ICU admissions to the level of a commissioning region, this assumes that each region behaves as a homogeneously allocated ‘pool’ of ICU beds, which is not necessarily true because inter-hospital ICU-to-ICU transfers may not be feasible for both operational and clinical reasons. We do not have more granular data available, but it is worth considering the likelihood that individual hospitals will reach critical capacity before the whole region. This has already been seen within the pandemic at certain London hospitals [19]. In this sense our predictions represent a ‘best case’ scenario and cannot be used for decision making at the level of an individual unit.

Building on this theme, we forecast the percentage of COVID-19 bed requirements in isolation, figures which do not reflect competing ICU burden. Since UK bed occupancy is typically greater than 80% and may frequently exceed 100% [7], clearly not all beds can simply be re-allocated for COVID-19 patients. This may be especially true for specialist ICUs (e.g. neuroscience or cardiac) which may not be able to entirely reconfigure, or high-dependency beds which may not be able to provide mechanical ventilation routinely. Furthermore, we assumed that all adult critical care bed spaces can be used for mechanically ventilated ICU patients, which operationally may not be possible for a variety of reasons including equipment and staff availability. Thus, the precise percentage of additional COVID-19 patients that will actually exhaust routine capacity will vary from unit to unit, particularly in ICUs with a substantial post-operative elective surgical workload. Finally, front-line clinicians reviewing our results in these early days of the pandemic will highlight the disconnect between our estimated ‘confirmed’ cases and the substantial operational workload that is being created by treating all suspicious cases of respiratory failure as COVID-19 until proven otherwise.

At the time of writing, the potential challenge that faces the health system (including intensive care) is essentially without precedent and has resulted in social and political interventions that have never been seen in peacetime. As such, there is little relevant precedent to judge the utility of approaches such as ours against. We hope that regardless of the assumptions inherent within our model (which we feel are based on best available data) that the overarching message of the need for urgent surge capacity is timely. Finally, we hope that our transparency in releasing our code, model assumptions, and results in a dynamic web format embodies a new paradigm in rapidly developed and reviewed science that persists beyond the current pandemic.

## Conclusions

Early warning of an impending need for ICU surge capacity is crucial if there is to be sufficient time to re-configure services. We have shown that ultra-early data can be used to make time-sensitive forecasts of ICU occupancy. We show, subject to our assumptions, that it is credible that ICU requirements may become challenging within weeks. There remains a significant degree of uncertainty in the predictions due both to limitations of the reporting data and modelling assumptions. This emphasises the need for the collection of real-time patient-facing local data by initiatives such as CHESS [18] and a dynamic approach to improving models as new data becomes available.

## Data Availability

Source data is already publicly available.

https://zenodo.org/badge/latestdoi/131309845

## Acknowledgments

The authors would like to thank Ronan O’Leary, Isobel Ramsay, David Menon and Tom Borchert for useful discussions as well as Mark Cresham for rapidly procuring computer facilities for our online model. Additionally we would like to thank Chris Fryer for crowd-sourced peer review of the manuscript and code which have been invaluable.

